# COVID-SAFER: Deprescribing Guidance for Nirmatrelvir-ritonavir Drug Interactions in Older Adults

**DOI:** 10.1101/2022.03.01.22271254

**Authors:** Sydney B. Ross, Émilie Bortolussi-Courval, Ryan Hanula, Todd C. Lee, Marnie Goodwin Wilson, Emily G. McDonald

## Abstract

**Importance:** Older adults, at high-risk of developing complications from COVID-19, could benefit from nirmatrelvir-ritonavir, an oral antiviral treatment for outpatients at high risk of complications from COVID-19; however, due to its potent CYP3A4 inhibition, nirmatrelvir-ritonavir is associated with many drug-drug interactions (DDI).

**Objectives:** Identify how common DDIs are between nirmatrelvir-ritonavir, common medications, and PIMs in older adults with polypharmacy. Craft anticipatory deprescribing guidance for PIMs that interact with nirmatrelvir-ritonavir to help prioritize deprescribing resources, and increase the proportion of older adults potentially benefitting from treatment.

**Design:** In this secondary analysis, we retrospectively analyzed all patients in the MedSafer cluster randomized deprescribing trial (N=5698 participants) to investigate the proportion of older adults (age >65) with polypharmacy (≥5 usual home medications) who would be ineligible for treatment with nirmatrelvir-ritonavir due to pre-existing DDIs.

**Setting:** The setting of the primary study was in medical inpatient units at 11 Canadian acute care hospitals.

**Participants:** Hospitalized persons, age 65 years and older, on 5 or more daily home medications, with an expected survival of 3 months or longer were included in this secondary analysis.

**Main outcomes and measures:** We identified the prevalence of (PIMs), as defined by the MedSafer software. We then developed deprescribing guidance, so clinicians could proactively deprescribe in an effort to increase the proportion of older adults eligible for safe treatment with nirmatrelvir-ritonavir in the event of a SARS-CoV-2 infection.

**Results:** Of 5698 participants, a total of 3869 (68%) were taking a medication with a known nirmatrelvir-ritonavir DDI, and of these 823 (21%) had at least one PIM. Of 823 PIMs, 627 (76%) were medications with a known high risk DDI and 213 (26%) were considered moderate risk DDIs with nirmatrelvir-ritonavir. Many of the PIMs required “advanced deprescribing” and could not simply be stopped, held, or adjusted at the time of nirmatrelvir-ritonavir receipt.

**Conclusions and relevance:** Older adults are at high risk of developing severe complications from COVID-19. Deprescribing PIMs in advance of a COVID-19 infection could increase the proportion of older adults who can safely receive nirmatrelvir-ritonavir, in addition to the usual benefits observed with medication management.

**Impact Statement:** We certify that this work is novel. This timely clinical investigation explores the unforeseen consequences of polypharmacy and the use of potentially inappropriate medication in older adults during the COVID-19 pandemic. This manuscript addresses the many drug-drug interactions between nirmatrelvir-ritonavir, an antiviral treatment for COVID-19, and potentially inappropriate medications in older adults with polypharmacy from the MedSafer cluster randomized trial. Our work highlights that the pandemic has created an even greater urgency to examine the medication lists of older adults and proactively deprescribe to improve the safety and tolerability of different COVID-19 treatments.

**Key Points:**

- **Question:** How does polypharmacy affect the eligibility of older adults to receive Nirmatrelvir-ritonavir?
- **Findings:** 68% of older adults in the MedSafer cluster randomized trial had a DDI with nirmatrelvir-ritonavir, and 21% were taking at least 1 potentially inappropriate medication.
- **Meaning:** Due to its potent CYP3A4 inhibition, nirmatrelvir-ritonavir is associated with many drug-drug interactions (DDI).

## Introduction

Since the beginning of the pandemic, hundreds of thousands of older Americans have died from complications of COVID-19.^[1 2]^ Despite the availability of severe acute respiratory syndrome coronavirus 2 (SARS-CoV-2) vaccination, older adults remain one of the most vulnerable populations in terms of hospitalization and death due to COVID-19.^[3]^ Most recently, there has been a resurgence in cases brought on by new variants such as Omicron.^[4]^ Exponential spread, viral mutations, and waning of immunity support the need for both an agile vaccination strategy and an ongoing urgency to develop and test effective therapies.^[4 5]^ On December 22nd, 2021, the FDA granted an emergency use authorization (EUA) for nirmatrelvir-ritonavir (marketed as Paxlovid), the first oral antiviral for the treatment of COVID-19^[6 7]^. Nirmatrelvir-ritonavir contains co-packaged tablets of nirmatrelvir (2×150 mg) and ritonavir (100 mg) and is indicated for the outpatient treatment of mild-to-moderate COVID-19 in adults and pediatric patients (12 years of age and older weighing at least 40 kg) with positive results of direct SARS-CoV-2 viral testing, and who are at high risk for progression to severe COVID-19, including hospitalization or death.^[8]^

An initial randomized clinical trial of nirmatrelvir-ritonavir demonstrated a reduction in risk of all-cause hospitalization or death by 88% (95% CI [75% -94%]) compared to placebo in high risk, symptomatic, unvaccinated adults with COVID-19.^[6 7]^ Comparatively, it is one of the most effective treatments for COVID-19 outpatients, among a number of novel or repurposed medications already studied.^[9 10]^

During the first wave of the pandemic, one such repurposed medication (hydroxychloroquine) generated significant interest, but ultimately proved to be ineffective in randomized controlled trials.^[11]^ However, hydroxychloroquine shed light on an important issue: many older adults could not safely receive this medication or enroll in clinical trials because of important pre-existing drug-drug interactions (DDIs).^[12]^ Our team conducted an analysis where we theoretically exposed a cohort of 1001 hospitalized older adults to hydroxychloroquine, and found that ∼60% of patients had potential DDIs and 43.2% of these involved so-called potentially inappropriate medications (PIMs). ^[12]^ With such a large proportion of patients identified with DDIs due to PIMs and polypharmacy, we called for the rationalization of medication lists in older adults with a focus on medication optimization and proactive deprescribing.

Millions of treatment courses of nirmatrelvir-ritonavir have now been purchased worldwide. Nirmatrelvir-ritonavir also has a long list of DDIs, limiting its safe use, especially among older adults with polypharmacy.^[10]^ Even with short term use, the ritonavir component strongly inhibits CYP3A4, an important enzyme for the metabolism of many drugs. It therefore has the potential to alter serum concentrations and therefore lead to toxicity or reduced efficacy of several common medications.^[10 13]^

Our aims for the present study were two-fold. The first was to identify how common DDIs are between nirmatrelvir-ritonavir, common medications, and PIMs in older adults with polypharmacy. We hypothesized that many patients would not be able to safely receive this oral antiviral. The second aim was to craft anticipatory deprescribing guidance for PIMs that interact with nirmatrelvir-ritonavir to help prioritize deprescribing resources, and ultimately, to increase the proportion of older adults who could benefit from treatment.

## Methods

The first COVID-SAFER study was conducted with hydroxychloroquine, using data from the pilot MedSafer study.^[12]^ For this COVID-SAFER: Nirmatrelvir-ritonavir study, we analyzed data from the 5698 patient cluster randomized MedSafer trial, which ran from August 2017 to January 2020.^[14]^ Briefly, inclusion criteria identified patients 65-years and older, taking five or more medications, admitted to one of the 11 study hospitals. Data pertaining to medications and comorbidities were analyzed with the MedSafer deprescribing software. The software applies a ruleset for the identification of PIMs which includes common recommendations from expert groups.^[15-17]^ MedSafer generates a prioritized, individualized deprescribing report according to the perceived level of medication harm (high, intermediate, or low risk/little added value), and provides the rationale for deprescribing with appropriate tapering protocols where required.^[18]^ The MedSafer trial compared the provision of individualized deprescribing opportunity reports to usual care for the primary outcome of prevention of post-discharge adverse drug events (ADEs) and, among several secondary outcomes, the augmentation of deprescribing of PIMs at hospital discharge.

For the present study, we utilized the data from the MedSafer trial as a representative cohort of older adults with polypharmacy. Over 85% of the patients in the MedSafer study were community-dwelling and all the participants had risk factors for developing severe complications from COVID-19, by virtue of age and comorbidity.

First, we searched the literature for medications with known DDIs with nirmatrelvir-ritonavir by examining the product monograph,^[8]^ referring to DDI websites and other publications,^[19]^ and reviewing the exclusion criteria for the randomized controlled trial which led to the EUA (EPIC-HR; NCT04960202).^[20]^ Medications with known interactions were grouped according to the American Hospital Formulary Service (AHFS) classification^[21]^ (Table 1) and divided into two categories: chronic medications and medications typically prescribed for a short course (e.g., antibiotics). We theoretically “exposed” the MedSafer trial cohort to treatment with nirmatrelvir-ritonavir (twice daily for 5 days), identified DDIs, and linked these with the associated potentially harmful outcome, such as medication toxicity, cardiac arrhythmias, increased risk of bleeding, respiratory depression, or death. We classified the DDIs as either severe (e.g., cardiac arrhythmias and respiratory depression) moderate (e.g., increased therapeutic drug level/risk of toxicity) or other (e.g., reduced effectiveness of the drug and/or reduced effectiveness of nirmatrelvir-ritonavir). We further identified the risk of an ADE requiring closer monitoring during therapy (e.g., anti-coagulation parameters or behavioral symptoms; Supplemental Table 1).

Next, we applied the MedSafer rules to determine the proportion of patients receiving one or more PIMs interacting with nirmatrelvir-ritonavir. When present, we also identified the triggering condition associated with each PIM (e.g., orthostatic hypotension, dementia, delirium, renal failure). Finally, we crafted anticipatory deprescribing guidance to avoid potential DDIs. Recommendations were based on the literature and the expert opinion of the authors.^[15-17 19]^ Ideally, we considered a scenario wherein PIMs were proactively deprescribed weeks to months in advance of ever requiring treatment with nirmatrelvir-ritonavir. As we are aware that such an approach is not always feasible, we also crafted mitigation strategies that included: do not co-administer, hold medication, adjust dose, and/or monitor clinically, based on a prior publication by the authors (Supplemental Table 1).^[12 19]^

**Table 1.** Common Medications with Potential Drug-Drug Interactions with Nirmatrelvir-ritonavir

| Interacting Drug Class | Drug Name | Severity | Potential interaction with Nirmatrelvir-ritonavir |
| --- | --- | --- | --- |
| Antiarrhythmics | Quinidine<br>Flecainide<br>Propafenone<br>Amiodarone<br>Dronedarone | Severe | CYP3A4 Inhibition may increase the serum concentrations. Potential for serious and/or life-threatening reactions, such as cardiac arrhythmias. |
|  | Digoxin | Severe | P-gp substrate inhibition may increase the serum concentration of digoxin, increasing toxicity. |
|  | Ranolazine | Severe | Potential for serious and/or life-threatening reactions. |
| Oral antithrombotic agents | Warfarin | Moderate | CYP3A4 Inhibition may decrease serum concentrations<br>Potential for reduced anticoagulation. |
|  | Dabigatran | Moderate | Mixed P-gp substrate inhibition and induction. Increased risk of bleeding, and possible toxicity. |
|  | Rivaroxaban | Severe | CYP3A4 and P-gp substrate Inhibition may increase the serum concentrations, increasing toxicity and risk of bleeding. |
|  | Apixaban | Moderate |  |
|  | Edoxaban | Moderate | P-gp inhibition may increase plasma concentrations. |
|  | Ticagrelor | Severe | CYP3A4 inhibition may increase plasma concentrations. |
|  | Clopidogrel | Severe | CYP3A4 induction, decreased plasma concentration.<br>Increased risk of atherothrombotic events. |
| Anticonvulsants | Phenobarbital<br>Carbamazepine<br>Phenytoin | Other | CYP3A4 induction, decreased plasma concentration and reduced clinical effects of nirmatrelvir-ritonavir. |
| Lipid Modifying agents | Lovastatin<br>Simvastatin | Severe | CYP3A4 inhibition may increase serum concentrations.<br>Potential for serious myopathy or rhabdomyolysis. |
|  | Atorvastatin,<br>Rosuvastatin | Moderate |  |
| PDE5i | Sildenafil<br>Vardenafil | Severe | CYP3A4 inhibition may increase serum concentrations.<br>Potential for serious hypotension or syncope. |
| Alpha1-blocker | Alfuzosin | Severe | CYP3A4 inhibition may increase serum concentrations.<br>Potential for serious reactions, such as hypotension. |
|  | Tamsulosin | Moderate |  |
| Antipsychotics | Clozapine<br>Lurasidone<br>Quetiapine<br>Pimozide | Severe | CYP3A4 inhibition may increase serum concentrations. Potential for serious and/or life-threatening reactions, such as cardiac arrhythmias, hematologic abnormalities, and coma. |
| NSAIDs | Piroxicam | Severe | Potential for life-threatening cardiac arrhythmias. |
| GI Motility | Cisapride | Severe | Potential for life-threatening cardiac arrhythmias. |
| Opioids | Fentanyl | Severe | CYP3A4 inhibition, increased serum concentrations. Serious/life-threatening respiratory depression. |
|  | Methadone | Other | CYP3A4 induction, decreased plasma concentration and reduced clinical effect of methadone. |
| Immuno-suppressants | Tacrolimus | Severe | CYP3A4 inhibition may increase plasma concentrations. Substantial risk for rapid onset of tacrolimus toxicity. |
| Ergot Derivatives | Ergotamine<br>Ergonovine | Severe | CYP3A4 inhibition may increase serum concentrations. Potential for serious and/or life-threatening reactions. |
| Sedative hypnotics | Sedative hypnotics | Severe | CYP3A4 inhibition may increase serum concentrations. Potential for extreme sedation/respiratory depression. |
| Anti-gout | Colchicine | Severe | CYP3A4 inhibition may increase serum concentrations. Potential for serious and/or life-threatening toxicity. |

\* Many of these medications are potentially inappropriate only under certain clinical circumstances (please refer to Table 2 for potential triggering conditions); **potential inappropriate medications are indicated in bold.**
PIM= potentially inappropriate medication; CNS= central nervous system; NSAIDs=nonsteroidal anti-inflammatories; PDE5i=phosphodiesterase-5-inhibitors.
Not listed in this table are the following medications with important drug-drug interactions with nirmatrelvir-ritonavir: estrogen containing hormone contraceptives; chemotherapies; antifungals (e.g., voriconazole) and antimycobacterials (e.g., rifampin); long-acting inhaled beta agonists (e.g., salmeterol) as these were not collected in the MedSafer study.

Of note, we did not account for DDIs with over the counter or naturopathic medications. The most important one being St. John’s Wort, which cannot be co-administered with nirmatrelvir-ritonavir, as it may lead to loss of virologic response and the possible development of viral resistance.^[22 23]^ Furthermore, as MedSafer did not capture most systemic chemotherapies, these were not included, but are potentially relevant.

## Results

The MedSafer^[18]^ trial enrolled 5698 patients 65 years and older who were taking 5 or more medications and survived to hospital discharge. The median age of the cohort was 78 years (IQR 71-86) and 50.2% or participants were female. The most prevalent comorbidities were hypertension (74%), type 2 diabetes (39%), ischemic heart disease (34%), and atrial fibrillation (32%). The median number of daily at-home medications was 10 (IQR 8-14), with a median of 2 PIMs (IQR 1-4) identified per participant.

Of the 5698 patients, 3869 (68%) were prescribed one or more usual home medications that could interact with nirmatrelvir-ritonavir. The most commonly prescribed medications with known DDIs were oral antithrombotic medications (e.g., apixaban, warfarin, clopidogrel; 2131/5698 or 37%), statins (e.g., atorvastatin; 1901/5698 or 33%), and alpha1-adrenoceptor antagonists (e.g., alfuzosin, tamsulosin; 779/5698 or 14%) (Table 2). From those with DDIs with nirmatrelvir-ritonavir, MedSafer identified 823/3869 (21%) patients as receiving 1 or more PIMs: 627/823 (76%) with one or more high risk interacting PIMs and 213/823 (26%) with one or more moderate risk interacting PIM. The most common cause of DDIs identified was CYP3A4 inhibition by nirmatrelvir-ritonavir in 34/49 (69%) of the DDIs.

**Table 2.**
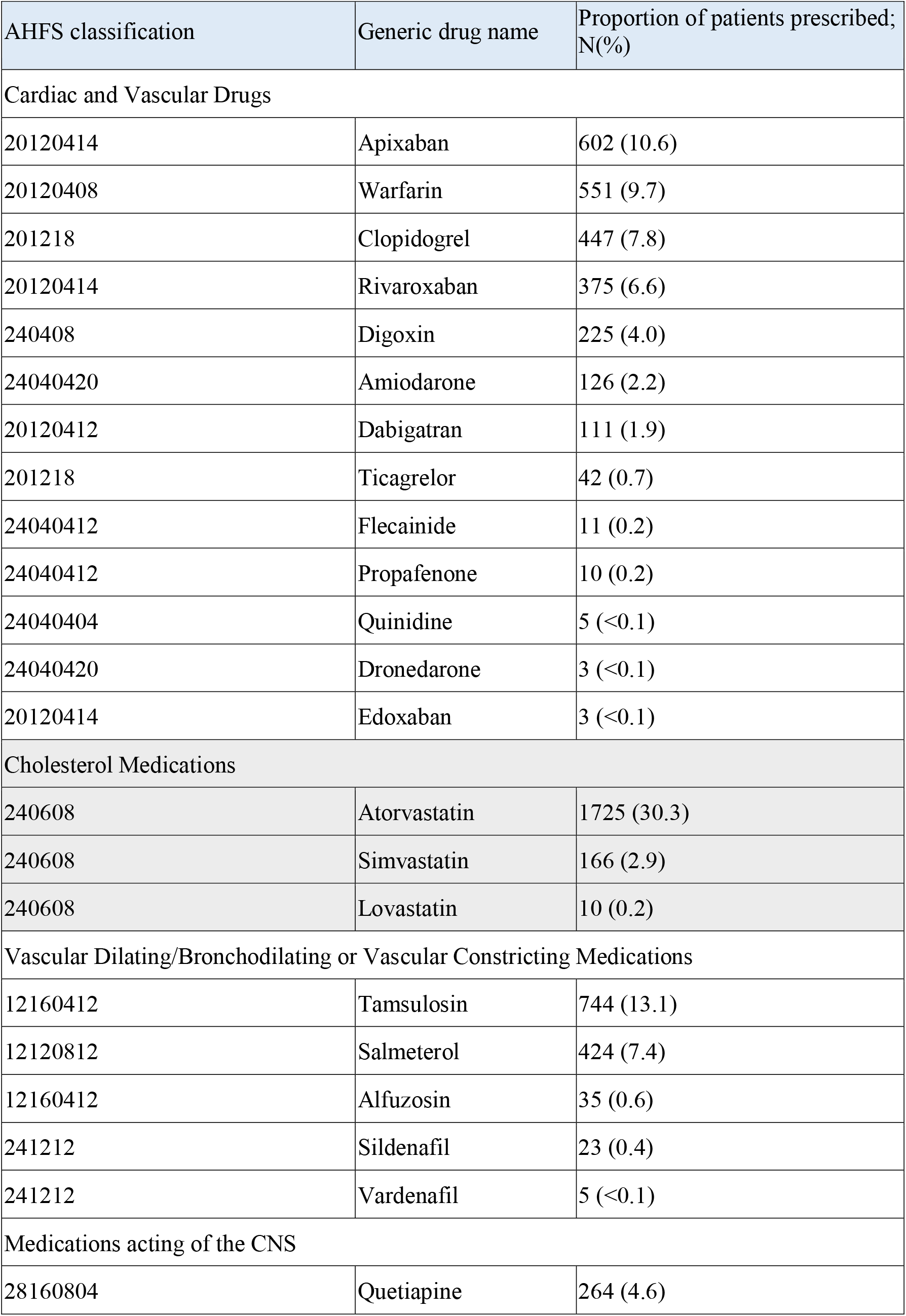
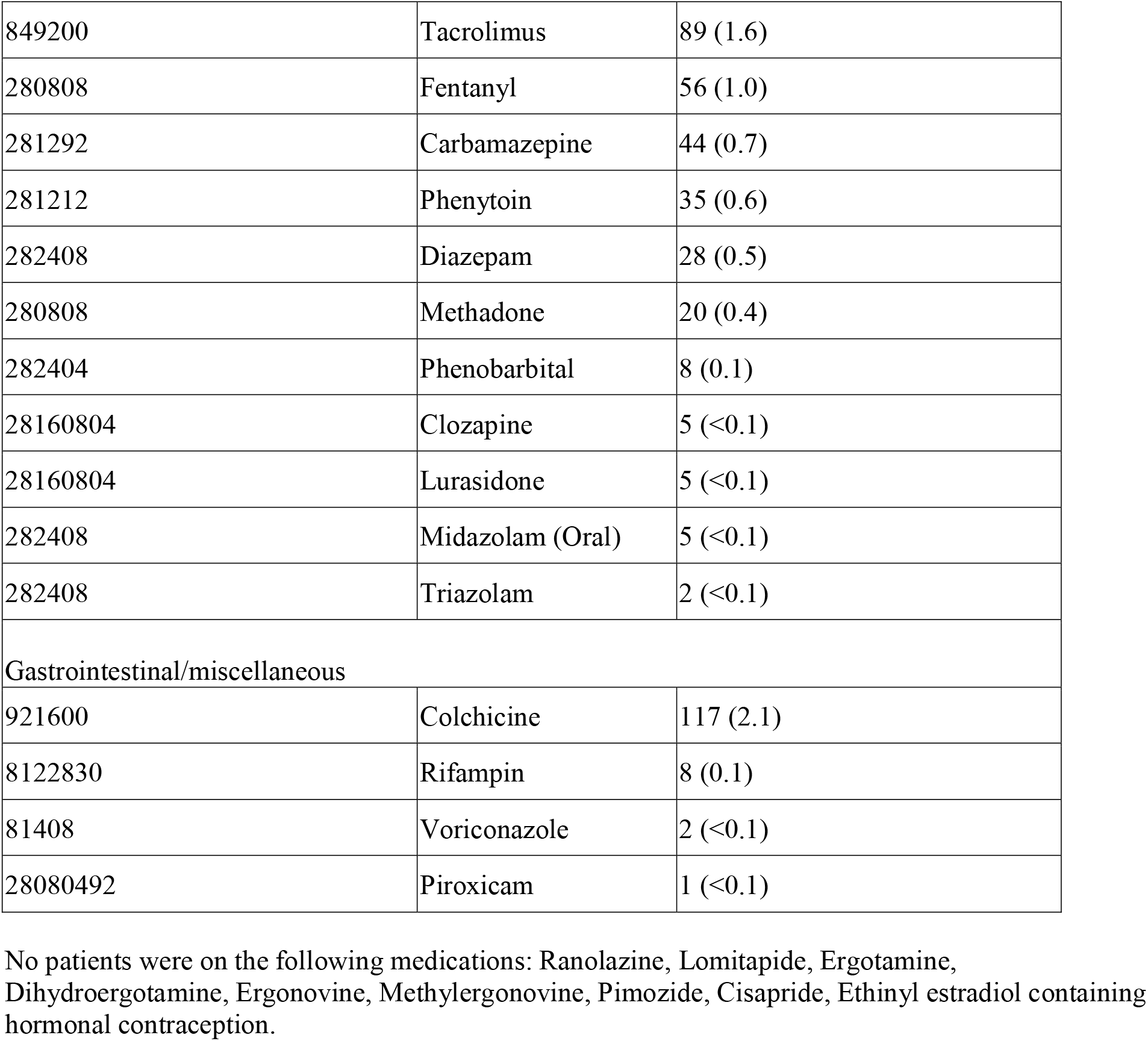
Prevalence of Drug-Drug interactions with Hypothetical Nirmatrelvir-ritonavir Exposure

| AHFS classification | Generic drug name | Proportion of patients prescribed; N(%) |
| --- | --- | --- |
| Cardiac and Vascular Drugs |  |  |
| 20120414 | Apixaban | 602 (10.6) |
| 20120408 | Warfarin | 551 (9.7) |
| 201218 | Clopidogrel | 447 (7.8) |
| 20120414 | Rivaroxaban | 375 (6.6) |
| 240408 | Digoxin | 225 (4.0) |
| 24040420 | Amiodarone | 126 (2.2) |
| 20120412 | Dabigatran | 111 (1.9) |
| 201218 | Ticagrelor | 42 (0.7) |
| 24040412 | Flecainide | 11 (0.2) |
| 24040412 | Propafenone | 10 (0.2) |
| 24040404 | Quinidine | 5 (<0.1) |
| 24040420 | Dronedarone | 3 (<0.1) |
| 20120414 | Edoxaban | 3 (<0.1) |
| Cholesterol Medications |  |  |
| 240608 | Atorvastatin | 1725 (30.3) |
| 240608 | Simvastatin | 166 (2.9) |
| 240608 | Lovastatin | 10 (0.2) |
| Vascular Dilating/Bronchodilating or Vascular Constricting Medications |  |  |
| 12160412 | Tamsulosin | 744 (13.1) |
| 12120812 | Salmeterol | 424 (7.4) |
| 12160412 | Alfuzosin | 35 (0.6) |
| 241212 | Sildenafil | 23 (0.4) |
| 241212 | Vardenafil | 5 (<0.1) |
| Medications acting of the CNS |  |  |
| 28160804 | Quetiapine | 264 (4.6) |
| 849200 | Tacrolimus | 89 (1.6) |
| 280808 | Fentanyl | 56 (1.0) |
| 281292 | Carbamazepine | 44 (0.7) |
| 281212 | Phenytoin | 35 (0.6) |
| 282408 | Diazepam | 28 (0.5) |
| 280808 | Methadone | 20 (0.4) |
| 282404 | Phenobarbital | 8 (0.1) |
| 28160804 | Clozapine | 5 (<0.1) |
| 28160804 | Lurasidone | 5 (<0.1) |
| 282408 | Midazolam (Oral) | 5 (<0.1) |
| 282408 | Triazolam | 2 (<0.1) |
| Gastrointestinal/miscellaneous |  |  |
| 921600 | Colchicine | 117 (2.1) |
| 8122830 | Rifampin | 8 (0.1) |
| 81408 | Voriconazole | 2 (<0.1) |
| 28080492 | Piroxicam | 1 (<0.1) |

No patients were on the following medications: Ranolazine, Lomitapide, Ergotamine, Dihydroergotamine, Ergonovine, Methylergonovine, Pimozide, Cisapride, Ethinyl estradiol containing hormonal contraception.

Some common deprescribing opportunities for PIMs included: 1) statins in older individuals with limited life expectancy, 2) antipsychotics for sleep or agitation, 3) opioids for chronic non-cancer pain and 4) alpha-1 blockers in patients predisposed to orthostatic hypotension (Supplemental Table S2).

## Discussion

In a representative trial population of older adults with polypharmacy, we found that 68% of older adults were taking one or more medications with a DDI with nirmatrelvir-ritonavir and 21% were prescribed one or more interacting PIMs. Many of these (two thirds) were high risk interactions and some would require advanced deprescribing to allow for treatment with nirmatrelvir-ritonavir (the most obvious example being amiodarone due to its extremely long half-life). In the absence of an advanced plan to deprescribe, many older adults in the cohort would require an alternative COVID-19 therapy because the interactions could not be mitigated by holding the interacting medication.

These findings build on our previous research, calling for proactive deprescribing of PIMs among older adults.^[12]^ This would not only reduce the general burden of overmedication commonly observed among 40-50% of older adults,^[24]^ but would also potentially render more people eligible to receive certain COVID-19 therapies. This motivated our secondary study aim which was to craft deprescribing guidance for PIMs that interact with nirmatrelvir-ritonavir to provide clinicians with a practical tool, to help triage deprescribing resources, and to highlight PIMs that should be addressed as soon as possible during the pandemic.

The most common high risk interacting PIMs were lipid lowering agents being given for primary prevention in patients with a limited life expectancy.^[25]^ Deprescribing statins in this population has the added benefit of improving quality of life.^[25]^ Also identified were combination antithrombotics/anticoagulants, alfuzosin and tamsulosin in people predisposed to orthostatic hypotension, atypical antipsychotics prescribed for sleep or agitation, and chronic opioids for non-malignancy associated pain. The added benefit of deprescribing these classes of medications is the potential to reduce medication related side effects and harmful ADEs such as bleeding,^[26]^ falls,^[27]^ and fractures.^[28]^ Of note, opioids for chronic non-cancer pain need to be deprescribed well in advance in order to administer nirmatrelvir-ritonavir due to the risk of acute opioid withdrawal in the absence of tapering.

Limitations of this study are the theoretical nature of the “drug exposure” to only one treatment for COVID-19; however, with millions of courses of nirmitrelvir-ritonavir purchased worldwide, it is expected to be a common exposure. This analysis could also be replicated for other promising potential outpatient treatments, such as fluvoxamine, which also has several DDIs.^[29]^ We also focused this paper on PIMs; however, the majority of interacting medications were not PIMs. We have provided medication management guidance for many of these medications elsewhere.^[12]^ It is also important to recognize that stopping PIMs requires clinical judgment when formulating a deprescribing plan. The strength of this study is the large sample size of representative community dwelling older adults with polypharmacy who are at high risk of severe COVID-19 complications; arguably, one of the populations most likely to benefit from treatment with nirmatrevir-ritonavir.

## Conclusion

In this cohort, many older adults were prescribed chronic PIMs that had high risk DDIs with nirmatrelvir-ritonavir. Some of these would need to be stopped well in advance of treatment. We suggest clinicians look to proactively deprescribe PIMs to allow older adult populations to safely benefit from the receipt of promising emerging treatments against COVID-19.

## Data Availability

All data produced in the present study are available upon reasonable request to the authors.

## Acknowledgments

We wish to thank the patients who enrolled in the MedSafer study who agreed to participate in research into polypharmacy.

## Author contributions

(study concept and design: EGM, TCL, SBR, acquisition of subjects and/or data: EGM, TCL, analysis and interpretation of data: MGW, preparation of manuscript: all authors, revision of the manuscripts: all authors).

## Information on author access to data

authors had access to the anonymized primary data from the original MedSafer study, used strictly for the purposes of the primary study and the present secondary analysis.

## Financial Disclosure

This project did not receive any funding.

## Conflict of interest

Todd Lee and Emily McDonald have received research funding from the Canadian Frailty Network, the Canadian Institutes for Health Research, and the Centre for Aging and Brain Health Innovation for the development of MedSafer. Drs Lee and McDonald, along with McGill University, own the intellectual property to MedSafer and are executives of MedSafer CORP, a for-profit company. Drs. Lee and McDonald receive research salary support from the Fonds de recherche du Québec - Santé.

## Sponsor’s role

This study was not funded.

## Disclaimers

None to disclose

## Information on previous presentation of the information reported in the manuscript

The MedSafer study^[18]^ contains the primary data from which the present study was derived.

## Notes

### Clinical Protocols

https://cdn.jamanetwork.com/ama/content_public/journal/intemed/0/ioi210080supp1_prod_1642198696.39131.pdf?Expires=1649169920&Signature=K6WZSgGHyo~r2cGwewqSrg5DIMoFzLp0FwPk0zfr4dpHX9i-PlVRbXA0Nuk8ImICsFbNHJEYy1KjsnGV4379-cPr0MaJE775EzI4XK7SM0OLugMEf-VPseYsUXhms154K5icDbcUXKIYsNIP~0hNbIeklGM1bjhaRWUv-Sfqs2jUqb5ZFnGA4xZ7eDogNGE0TbmG39kvGtCjZGYhKBIJKhYCNtIl9QTrlcczm~X0gzpuNw43wmarAFlezBEVtSb9vDB-F4GxlgcSAbqEEk5Kd1tNl1JYpIqMNSE5yHRBqWosiXck~XwloTYCWLGvgE4VIYbio1IpIxmuM1Ldo1LZ1g__&Key-Pair-Id=APKAIE5G5CRDK6RD3PGA

### Funding Statement

This study did not receive funding.

### Author Declarations

IRB of Foothills Medical Centre, Calgary, Alberta, Canada gave ethical approval for this work IRB of University of Alberta, Edmonton, Alberta, Canada gave ethical approval for this work IRB of St-Paul's Hospital (University of British Columbia), Vancouver, British Columbia, Canada gave approval for this work IRB of Kingston General Hospital, Kingston, Ontario, Canada gave approval for this work IRB of The Ottawa Hospital, Ottawa, Ontario, Canada gave approval for this work IRB of the University Health Network, Toronto, Ontario, Canada gave approval for this work IRB of McGill University Health Centre, Montreal, Quebec, Canada gave approval for this work Potentially eligible patients were approached for consent for a 30‐day post-discharge telephone interview. For patients lacking capacity, family or proxy provided consent as per Canadian ethics guidelines. Patients consented to have their data anonymized and used in secondary analyses from the initial MedSafer Study. Additional information on consent provision can be found here: https://jamanetwork.com/journals/jamainternalmedicine/article-abstract/2788297

