## Supplemental Tables S1 and S2 for "COVID-SAFER: Deprescribing Guidance for Nirmatrelvir-ritonavir Drug Interactions in Older Adults"

**Supplemental Table S1.** Medication Management for Potentially Inappropriate Medications with Nirmatrelvir-ritonavir Drug-Drug Interactions

| **AHFS Drug Class** | **Potentially Inappropriate Medication** | **Triggering Condition or Medication:** | **General Rationale for Deprescribing:** | **Preemptive Deprescribing Required (Y/N)** | **Suggested Medication management for Nirmatrelvir-ritonavir interaction IF unable to deprescribe in advance:** |
| --- | --- | --- | --- | --- | --- |
| 24040420 | Amiodarone | Atrial fibrillation | Amiodarone is associated with multiple toxicities including thyroid disease, pulmonary disorders, and QT- interval prolongation. Consider a safer alternative. | Y | DO NOT CO-ADMINISTER  If unable to deprescribe well in advance (weeks); likely precludes use of nirmatrelvir-ritonavir given increased risk of life-threatening cardiac arrhythmias. |
| 24040420 | Dronedarone | CHF | Potential to increase mortality in older adults with heart failure. | Y | DO NOT CO-ADMINISTER  If unable to deprescribe well in advance (weeks); likely precludes use of nirmatrelvir-ritonavir given increased risk of life-threatening cardiac arrhythmias. |
| 24040404  24040412 | Quinidine  Flecainide  Propafenone | Atrial fibrillation | Data suggest that rate control yields better balance of benefits and harms than pharmacological rhythm control for most older adults. | N | DO NOT CO-ADMINISTER  If unable to deprescribe or hold, likely precludes use of nirmatrelvir-ritonavir given increased risk of life-threatening cardiac arrhythmias. |
| 240408 | Digoxin | Atrial fibrillation, CHF, CKD | A higher dose of digoxin (>125mcg daily) has an increased risk of toxicity. | N | ADJUST DOSE  Exercise caution. Individualize dose or dosing schedule based on plasma concentration, renal function and treatment indication. If digoxin levels are not available, evaluate the risks/benefit of co-administration. |
| 20120408 | Warfarin | Flagged in combination with dual antiplatelet medications | Dual antithrombotic therapy increases the risk of major hemorrhage. | N | ADJUST DOSE  Closely monitor anticoagulation parameters (INR) during co-administration. Warfarin dose may need to be increased. |
| 20120412 | Dabigatran | Flagged in combination with dual antiplatelet medications | Dual antithrombotic therapy increases the risk of major hemorrhage. | N | ADJUST DOSE  Co-administration possible with dose reduction to 110 mg BID and clinical monitoring for signs of bleeding. |
| 20120414 | Rivaroxaban  Apixaban | Flagged in combination with dual antiplatelet medications | Dual antithrombotic therapy increases the risk of major hemorrhage. | N | DO NOT CO-ADMINSTER  Hold medication during co-administration with nirmatrelvir-ritonavir. Treatment should resume 3 days after last dose.  Rivaroxaban carries a very high risk for bleeding.  Apixaban carries a moderate risk of bleeding. Some product monographs indicate a lower dose (2.5 mg BID) may be co-administered. |
| 20120414 | Edoxaban | Flagged in combination with dual antiplatelet medications | Dual antithrombotic therapy increases the risk of major hemorrhage. | N | ADJUST DOSE  Co-administration is possible with dose reduction to 30 mg daily and clinical monitoring for signs of bleeding. |
| 201218 | Ticagrelor  Clopidogrel | Flagged in combination with dual antiplatelet medications; reduced life expectancy; palliative | Primary prevention with antiplatelet agents could be reconsidered in patients with limited life expectancy. | N | DO NOT CO-ADMINISTER; DRUG SUBSTITUTION  Prasugrel can be prescribed instead unless the patient's clinical condition contraindicates its use.  Co-administration with ticagrelor leads to increased risk of bleeding.  Co-administration with clopidogrel leads to reduced efficacy and risk of thrombosis. |
| 282404 | Phenobarbital | Flagged for all older adults who do not have a seizure disorder or psychiatric condition | High rate of physical dependence; tolerance to sleep benefits; risk of overdose at low dosages. | Y | DO NOT CO-ADMINISTER  If unable to deprescribe well in advance (weeks); precludes use of nirmatrelvir-ritonavir due to decreased efficacy of nirmatrelvir-ritonavir and risk of developing virologic resistance. |
| 240608 | Lovastatin  Simvastatin  Atorvastatin | Flagged for primary prevention in people with reduced life expectancy or receipt of palliative care | In patients with restricted life expectancy based on their age, demographics, and recent health utilization, statins may offer little benefit and possible harm. | N | HOLD  Hold medication during co-administration with nirmatrelvir-ritonavir.  Treatment should only resume 3 days after last dose.  Risk of severe myopathy and rhabdomyolysis. |
| 241212 | Sildenafil  Vardenafil | Flagged in combination with Isosorbide dinitrate | High risk of cardiovascular collapse. Avoid this combination of medications. | N | DO NOT CO-ADMINISTER  Hold medication during co-administration with nirmatrelvir-ritonavir. Treatment should resume 3 days after last dose. |
| 12160412 | Alfuzosin  Tamsulosin | Orthostatic hypotension | High risk of orthostatic hypotension; not recommended as routine treatment for hypertension. | N | DO NOT CO-ADMINISTER  Hold during therapy and monitor for emergence of symptoms.  Alfuzosin is absolutely contraindicated due to risk of severe orthostatic hypotension.  Tamsulosin carries a moderate risk. |
| 28160804  28160892 | Clozapine  Lurasidone  Quetiapine  Pimozide | Flagged for all older adults who do not have a psychiatric disorder such as schizophrenia or bipolar affective disorder | Don’t routinely use antipsychotics in parkinsonism, dementia and treatment of insomnia or sleep disorders. Increase the risk of stroke, falls, confusion, extrapyramidal side effects, aspiration, and death. Quetiapine and clozapine have a lower risk but in general their use should be avoided. | Y (for high doses) | DO NOT CO-ADMINISTER  If low dose for sleep or agitation (e.g., <25mg of quetiapine daily), consider holding during nirmatrelvir-ritonavir co-administration and monitor closely for emergence of behavioral symptoms.  For higher doses, do not co-administer. High risk of serious sedation and coma.  Deprescribing usually requires tapering for higher doses. |
| 28080492 | Piroxicam | Flagged for most older adults | Avoid NSAIDS with a history of ischemic heart disease, gastrointestinal hemorrhages, hypertension, gastritis, CKD, GERD, peptic ulcers as they may exacerbate the underlying condition. | N | DO NOT CO-ADMINISTER  Hold medication during co-administration due to potential for respiratory depression or hematologic abnormalities. |
| 280808 | Fentanyl | Flagged for all older adults who in the absence of cancer | Avoid opioids for chronic non-cancer related pain. | Y | DO NOT CO-ADMINISTER  Fatal respiratory depression can occur. temporarily holding chronic, regular use opioids is not advised as this can precipitate withdrawal in the absence of tapering. Deprescribing requires tapering. |
| 280808 | Methadone | Flagged for all older adults in the absence of cancer | Avoid opioids for chronic non-cancer related pain. | Y | ADJUST DOSE  Increased methadone dose may be necessary when co-administered. |
| 282408 | Diazepam  Midazolam (Oral)  Triazolam | Flagged for all older adults who do not have a seizure disorder or severe anxiety | In general, all benzodiazepines increase risk of cognitive impairment, delirium, falls, fractures, and motor vehicle accidents in older adults. | Y | DO NOT CO-ADMINISTER  If unable to deprescribe well in advance (weeks); likely precludes use of nirmatrelvir-ritonavir. Deprescribing requires tapering. |
| 563200 | Cisapride | Flagged for all older adults | Can cause extrapyramidal effects, including tardive dyskinesia; risk may be greater in frail older adults and with prolonged exposure. | N | DO NOT CO-ADMINISTER  Hold medication during co-administration with nirmatrelvir-ritonavir due to known risk of life-threatening cardiac arrhythmias. |
| 921600 | Colchicine | Flagged for patients with a reduced eGFR | There is a risk of colchicine toxicity at reduced eGFR. Xanthine-oxidase inhibitors (e.g. allopurinol, febuxostat) are first choice for prophylactic drugs in gout. | N | DO NOT CO-ADMINISTER  This interaction can be deadly, especially in patients with renal and/or hepatic impairment. Life-threatening and fatal drug interactions have occurred with co-administration. |

CKD= chronic kidney disease; GERD= gastroesophageal reflux disease; AHFS=American Hospital Formulary Service
